# Microscale Multiplexed Antigen-Specific Antibody Fc Profiling for Point-of-Care Diagnosis of Tuberculosis

**DOI:** 10.1101/2025.11.15.25340307

**Authors:** Sarah M Ali, Sai Preetham Peddireddy, Asma Hashim, Neda Rafat, Abhipsa Panigrahi, Lynette Stone, Marwou de Kock, Jishnu Das, Cheryl L. Day, Aniruddh Sarkar

## Abstract

Accurate, affordable tuberculosis (TB) diagnostics that do not require sputum samples are urgently needed for TB control and elimination. Prior serological TB tests have failed due to low accuracy in diagnosing active TB (ATB) in endemic areas where baseline seropositivity, due to latent infection (LTBI) or vaccination, is common. We combined high-throughput sample-sparing antibody-omics with machine learning to profile Fab and Fc features of Mycobacterium tuberculosis (Mtb)-specific antibodies in plasma from adults with ATB or latent infection (LTBI). A five-feature Fc-centric signature dominated by enhanced FcγR3B binding, distinguished ATB from LTBI (AuRoC>0.9). To translate this signature for point-of-care use, we engineered a multimeric, enzyme-labelled FcγR3B probe and integrated it with a microscale silver-metallization assay on a low-cost platform, yielding a cellphone-based optical readout from <10 µL of blood. A further optimized three-feature signature measured on this platform correctly classified ATB from LTBI and endemic controls in two independent cohorts (AuRoC>0.9), exceeding World Health Organization (WHO) target levels for rapid TB triage tests. This work establishes Fc-centric antibody profiling as a viable biomarker discovery route and delivers a multiplexed, sample-sparing, inexpensive test suitable for deployment in resource-limited settings.

## Introduction

Tuberculosis (TB), caused by *Mycobacterium* tuberculosis (Mtb), remains the leading cause of death from a single infectious agent^1^. In 2024, approximately 1.25 million people died from TB primarily in the low to middle income countries (LMICs) where TB is endemic^2^. This is despite availability of effective TB treatments. Lack of accurate and rapid yet affordable diagnostics is a significant contributor to TB-related mortality as there is a persisting major diagnostic gap of an estimated 4.2 million undiagnosed TB cases^3^. Key challenges in TB diagnostics include difficulty in distinguishing the heterogenous spectrum of Mtb infection as well as difficulties in obtaining and processing sputum samples, needed for commonly used bacteriological TB tests such as sputum smear microscopy, bacteriological culture, and nucleic acid amplification tests (NAATs) (e.g. GeneXpert)^4^. Additionally, significant trade-offs exist between cost, complexity and accuracy of existing diagnostic methods.

The WHO End TB Strategy, a global plan to eliminate TB, has identified a critical need for a low-cost biomarker-based TB diagnostic that uses easily accessible non-sputum samples, and which can be deployed at the point-of-care (POC) in resource-limited settings^2^. Antigen-based diagnostics such as the urine LAM test, are compatible with POC use but currently remain restricted to use only in cases of HIV/TB coinfection^5^. More sensitive versions of this test which may work for HIV-negative TB are under development^6^. For many infections, serological diagnostics are often deployed as primary screening tools as they offer a simple and inexpensive approach for detecting pathogen-specific antibodies (Abs) in easily accessible samples (e.g. a drop of blood). However, such Ab titer-based approaches have historically shown low and variable diagnostic accuracy, with especially poor specificity, in TB^7,8^. This is due to high levels of baseline Mtb-specific Ab seropositivity in TB-endemic regions. A major fraction of this is people with latent TB infection (LTBI) (e.g. >50% of the population in South Africa are estimated to have LTBI) who are seropositive but do not exhibit signs or symptoms of active disease^9^. Additionally, Mtb-specific Ab titers can be influenced by prior vaccination with Bacillus Calmette-Gúerin (BCG), the only currently licensed vaccine for TB. Attempts to define discriminative Ab titer cutoffs or to identify particular antigen-specificities to discriminate active TB (ATB) disease from LTBI, have also shown limited success^8^.

Emerging evidence suggests that Ab ‘quality’, or Fc functional properties of Abs, rather than quantity (titer) is a promising biomarker for delineating disease states and outcomes across infections, including for TB^10–13^. Ab Fc regions undergo rapid and dynamic diversification after infection, including alterations in glycosylation and Fc receptor binding. In TB, distinct Fc glycosylation patterns on Mtb-specific Abs have emerged as candidate biomarkers distinguishing ATB from LTBI^14–16^. Current antigen-specific Ab glycomics approaches involving approaches such as mass spectrometry (MS) and capillary electrophoresis (CE) however remain too expensive, sample-intensive and complex for use as rapid and inexpensive TB diagnostics in resource-poor settings.

In this work, we first develop and use a sample-sparing, highly multiplexed antibody-omics pipeline to comprehensively characterize antigen-specific Abs – both Fab and Fc regions – in serum/plasma samples, from individuals with ATB and those with LTBI. Coupled to machine-learning based analytics, this helped define a distinct Mtb-specific Ab Fc-based biomarker for ATB. We then developed a multimeric high-avidity Fc probe targeting this Fc modification which enabled its detection in an inexpensive POC format using probe-directed enzymatic metallization which provides a cellphone-based multiplexable optical readout. Finally, this biomarker and POC detection method was tested in two independent patient cohorts, verifying its ability to robustly detect ATB using a single drop (<10µL) of serum or plasma sample.

## Results

### Distinct Antibody-omic Profiles Emerge in ATB and LTBI

Antibody-omic profiling was carried out on plasma from a cohort of individuals **(Table. S1)** from South Africa (n=36, **Figure 1A)**, including those with ATB and LTBI. This highly multiplexed bead-based assay^17–19^ (see Methods for details) was employed to comprehensively assess both Fab and Fc Ab domain properties of Abs across the cohort^18^. Ab response was measured across multiple Mtb-specific (PPD, LAM, Ag85A, ESAT6, CFP10, HSPX, MPT64, MPT32, PSTS1 and GROES) and non-Mtb (Flu and Tetanus Toxoid) antigens. Measured features included Ab isotype and subclass [IgG (IgG1-IgG4), IgA (IgA1 and IgA2) and IgM], Fc receptor binding (FcγR1, FcγR2A, FcγR2B, FcγR3A and FcγR3B) and lectin-based glycosylation profiles (SNA for sialic acid; RCA for galactose). Thus, a high-dimensional dataset consisting of 160 Mtb-specific and 32 non-Mtb-specific coupled Ab Fab and Fc features for each individual **(Figure S1)** was collected.

**Figure 1.**
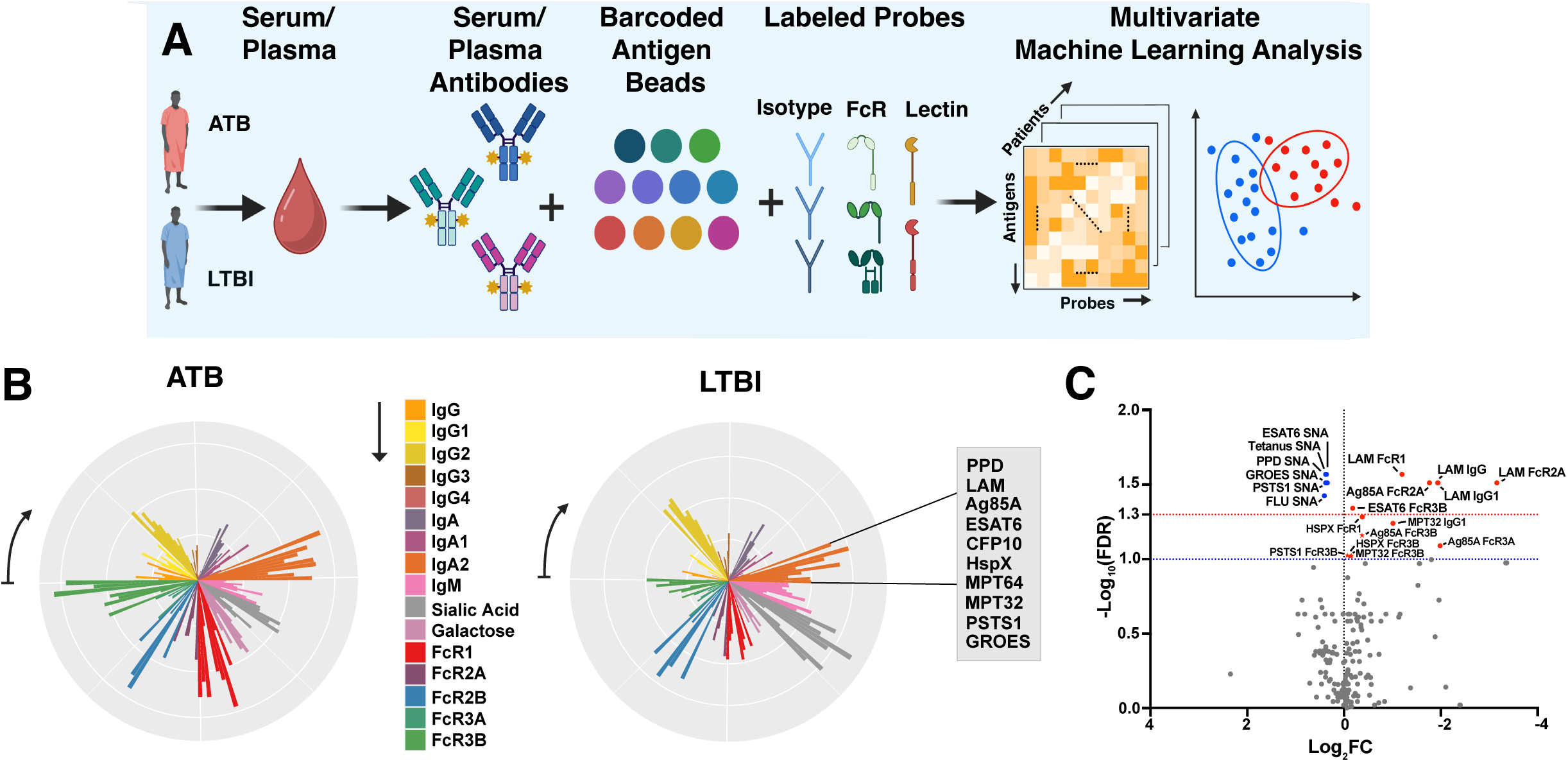
Multiplexed antigen-specific antibody Fab and Fc responses show differences in ATB (n=16) and LTBI plasma samples (n=20) from South African cohort. **(A)** Conceptual overview of the antibody-omics platform used to characterize and quantify plasma antibodies directed against Mtb and define ATB biomarkers **(B)** Visualization of measured ATB and LTBI Ab Fab and Fc features as petal plots. Lengths of radial bars represent min-max scaled medians of measured ATB and LTBI Ab features against Mtb antigens (n=160 total features). Each color, grouped in sectors, represents a single Fc feature, as shown, while Fab antigen-specificities are shown in each sector in the order clockwise as shown. **(C)** Volcano plot identifying the most discriminative features based on Mann-Whitney analyses with Benjamini-Hochberg correction for multiple testing (blue increased in LTBI; red increased in ATB).

Visualization of the multi-dimensional Mtb-specific Ab profiles, via the petal plots shown in **Figure 1B**, revealed striking differences, especially in FcγR binding (FcγR1: red bars, FcγR2A: purple bars, FcγR3B: green bars) and glycosylation (sialic acid: grey bars). ATB samples exhibited higher FcγR-binding, but lower sialic acid compared to LTBI samples. Interestingly, Mtb-specific IgG (light orange bars) & IgG1 (yellow bars) show differences as well with ATB samples showing higher levels than LTBI. Univariate analysis of the Mtb-specific and non-Mtb-specific Ab profiles (192 total features), shown in the volcano plot in **Figure 1C**, revealed significant differences (p<0.05, Mann-Whitney with Benjamini-Hochberg correction, red dotted line) in Mtb-specific Ab titers, Fcγ receptor binding and Ab glycosylation. ATB samples exhibited higher Mtb-specific IgG, IgG1 and FcγR1 (LAM), FcγR2A (LAM and Ag85A) and FcγR3B binding (ESAT6). LTBI samples showed higher sialylation of Abs across 5 Mtb and 2 non-Mtb antigens. We also noted differences in four additional Mtb-specific Ab FcγR3B binding features (red stars) as well (p <0.1, Mann-Whitney with Benjamini-Hochberg correction, blue dotted line) which warrant further investigation as, while they did not meet threshold for statistical significance, the direction of the effect was found to be consistent with the previous significant findings.

### Machine Learning Identifies Minimal Fab and Fc Ab Signature for ATB Diagnosis

A robust two-step machine-learning based method^17^ was then used to define a minimal set of Ab features as a biomarker for ATB. Feature selection using the least absolute shrinkage and selection operator (LASSO) was followed by classification with a support vector machines (SVM) model based on the down-selected features. This model was based on 5 features spanning both Ab isotype (Ag85A.IgM) and Fc (ESAT6.Sialic Acid, LAM.FcγR1, ESAT6.FcγR3B and HspX.FcγR3B) features **(Figure 2A)** and showed high diagnostic accuracy (AuRoC>0.96, **Figure 2B**). The model was also significantly predictive when measured in a k-fold cross-validation framework with permutation testing where permuted denotes the performance of the model on shuffled data in a matched cross-validation framework (negative control) **(Figure 2C)**. As an orthogonal way to visualize this stratification, we performed partial least squares discriminant analysis (PLS-DA) using these 5 down-selected features as well. This also demonstrated that these features could differentiate ATB and LTBI **(Figure 2D)**, with the relative contributions of features as shown in **Figure 2E**. Among the 5 selected features, all 3 FcγR features (LAM.FcγR1, ESAT6.FcγR3B and HspX.FcγR3B) were elevated in ATB while IgM and Ab sialyation were increased in LTBI.

**Figure 2.**
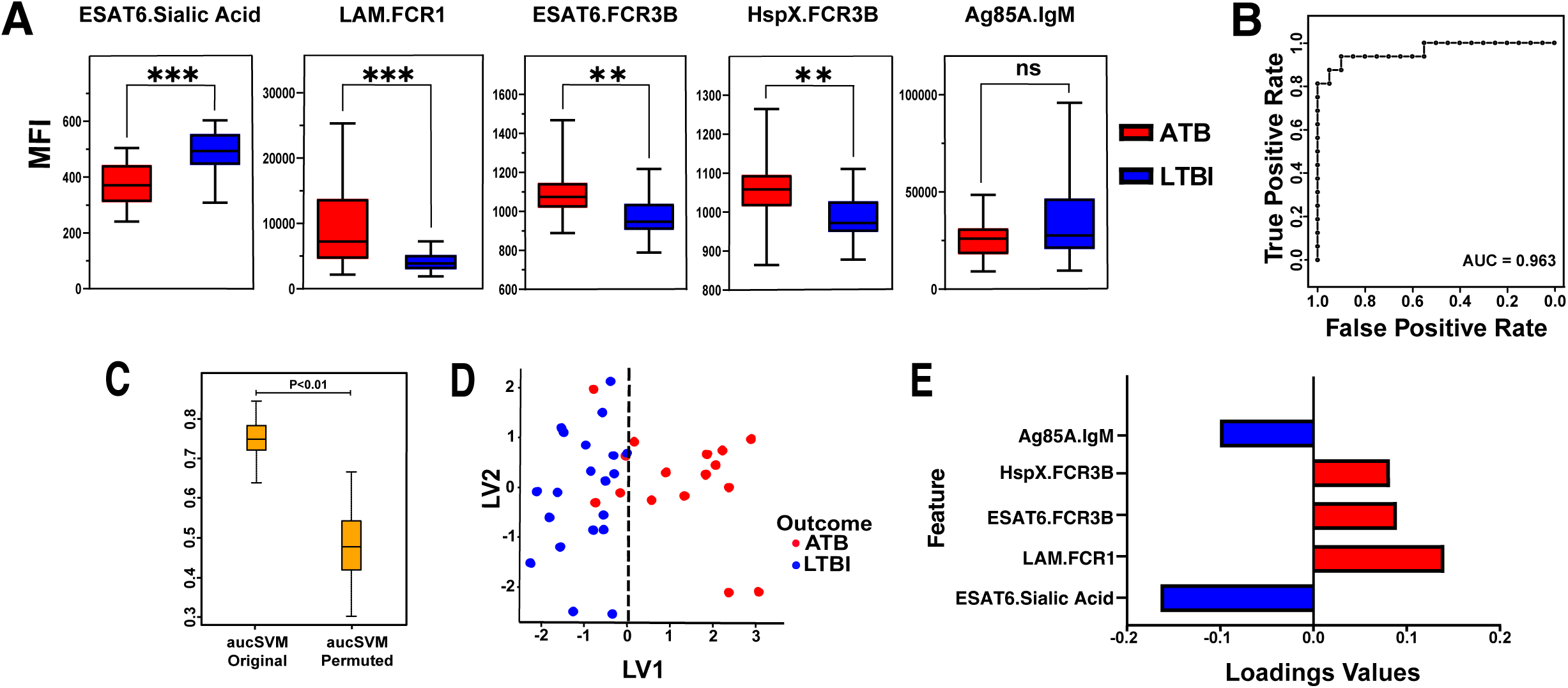
Machine-learning identifies minimal multivariate biomarker for ATB **(A)** Box and whisker plots of five LASSO-selected features in ATB (n=16) vs LTBI (n=20) in South African cohort. Statistical significance was assessed using Mann-Whitney test (***: p<0.001, **: p<0.01, ns: p>0.05). **(B)** Receiver Operating Curve (ROC) showing LASSO-SVM model performance using a combination of all five features in (A) to discriminate between ATB and LTBI samples. **(C)** Control plot showing performance of LASSO model on antibody-omic profiling data consisting of 192 Ab Fab + Fc features from South African cohort (n=36). Permuted denotes performance of the model on shuffled data in a matched cross-validation framework (negative control). **(D)** PLS-DA visualization using the five LASSO-selected features from the model in (B) to discriminate between ATB and LTBI. **(E)** PLS-DA loadings plot showing the relative importance of the five LASSO-selected features in discriminating between ATB and LTBI.

Overall, the above univariate and multivariate analyses revealed a role for Ab Fc properties taken together with Ab titer, measured across multiple Mtb antigens, as a biomarker for ATB. Two of the major discriminative Fc properties revealed by these analyses were FcγR3B binding and sialylation. Strikingly, higher FcγR3B binding to Abs across many Mtb antigens emerged here as a leading single Mtb-specific Fc difference in ATB in both univariate and multivariate analysis. Differences in Ab sialylation, while present across multiple Mtb-specific antigens as well, were also present in non-Mtb specific Abs (Flu and Tetanus toxoid) **(Figure 1C, Figure S2)**. Based on these findings, we selected multiplexed Mtb-specific Ab FcγR3B binding, along with Mtb-specific IgG titer for comparison, for translation to a POC format and further testing.

### Multimeric Fc Probe Coupled to Enzymatic Metallization Enables Fc-binding Measurement in a Multiplexable POC Platform

Two key challenges arise in developing an inexpensive and simple POC method to detect titer and Fcγ receptor binding-based biomarkers: first, achieving effective multiplexing with a straightforward, low-cost readout while maintaining ease of use; and second, overcoming the inherently low binding affinity of Fcγ receptors—particularly FcγR3B—which can typically limit assay sensitivity^20,21^.

Here, we overcome the first challenge by harnessing a microscale probe-directed enzymatic silver metallization method^22–25^. This converts analyte binding to localized enzymatically amplified silver substrate deposition to transduce biomarker concentrations into a visible, amplified layer of dry-stable silver metal. This enables simple and inexpensive multiplexed biomarker measurements, from a small sample volume, based on an optical density based quantitative readout which can be obtained using a cellphone camera and app or an inexpensive office scanner. Here we adapt this method to perform it on a flexible polymeric (laser-cut PDMS, see Methods for details) substrate, mounted on a typical microscope slide, for sensitive yet inexpensive detection of the multivariate Mtb-specific Ab biomarker **(Figure 3A).**

**Figure 3.**
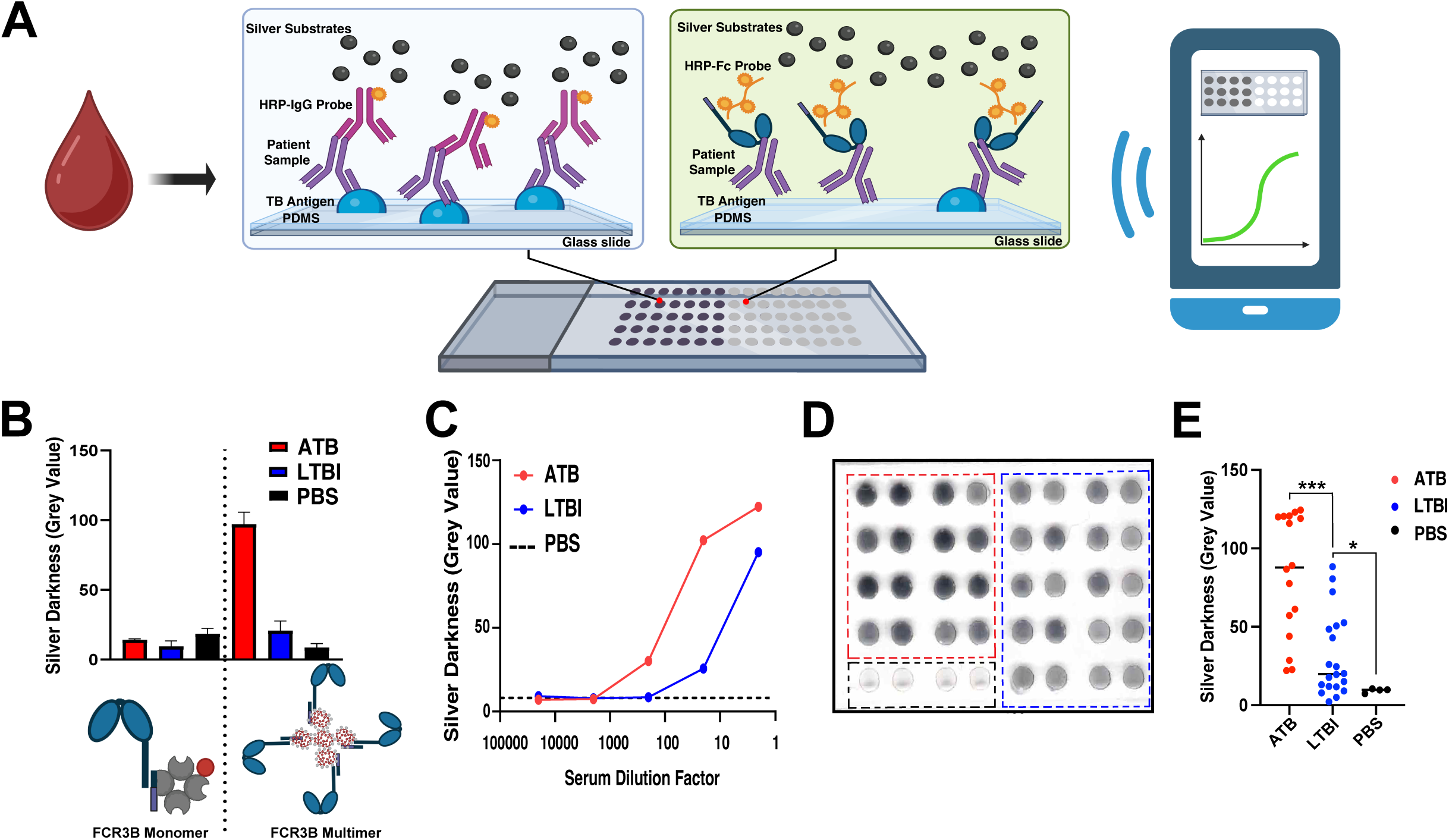
Development and validation of multimeric FcR3b probe-based biomarker assay on a point-of-care diagnostic platform. **(A)** Conceptual overview of enzymatic silver metallization-based measurement of IgG (in blue) and FcγR3B (in green) binding in plasma on flexible PDMS substrate mounted on a glass slide. **(B)** Evaluation of monomeric and multimeric enzyme-labeled FcγR3B probes **(C)** Quantification of silver darkness for detection of FcγR3B binding to Mtb Ag85A-specific antibodies from serial dilution of pooled ATB and LTBI plasma. **(D)** Silver metallization deposited in response to FcγR3B binding to Ag85A-specific antibodies in individual ATB (n=16) and LTBI plasma samples (n=20) from South African cohort and PBS control. **(E)** Quantification of silver metallization response of ATB and LTBI and PBS control shown in (D).

To overcome the challenge posed by Fcγ receptor binding affinity constraints, we took inspiration from the fact that in-vivo FcγR binding often relies on avidity through multimeric interactions^20^. Thus, to achieve high avidity binding to Mtb-specific Abs in-vitro, we developed a multimeric enzyme-labeled FcγR3B probe using biotinylated FcγR3B and a polymeric horseradish peroxidase (HRP). Evaluation of both monomeric **(Figure 3B, left)** and multimeric **(Figure 3B, right)** forms of the probe revealed that significantly improved binding signal on the POC platform was achieved using the multimeric probe. It also showed clear discrimination between ATB and LTBI samples across a range of pooled sample dilutions **(Figure 3C)**. **Figure 3D** shows a representative image of the POC assay, described above, using this multimeric probe, to measure FcγR3B binding to Mtb Ag85A-specific Abs, in individual ATB and LTBI samples from the South African cohort (n=36). **Figure 3E** shows the quantification of the darkness of these silver deposits. The differences in the silver spot darkness, between ATB and LTBI samples and a buffer control were clearly visible, and the quantification showed statistically significant differences between ATB and LTBI as well (p<0.001).

### Machine Learning Applied to POC Platform Readouts Identifies and Validates Minimal Titer-Fc**γ**R Signature for ATB Diagnosis Across Two Cohorts

Multimeric probe-enabled FcγR3B binding together with IgG titer levels were then evaluated on the POC platform across 5 different Mtb antigen specificities (Ag85A, LAM, PPD, ESAT6 and HspX) using ATB and LTBI plasma samples from South Africa (n=36) **(Figure S3A-B)**. The LASSO-SVM machine-learning pipeline, as described above, was applied next to the resulting 10-feature dataset to identify a minimal set of Ab features as a biomarker for ATB on the POC platform. The resulting 3-feature model **(Figure 4A)** consisting of titer (Ag85A.IgG) and FcγR3B-binding (HspX.FcγR3B and PPD.FcγR3B) features, demonstrated high diagnostic accuracy (AuRoC>0.9, **Figure 4B, in blue)**. The 3-feature model was still significantly predictive when measured in a k-fold cross-validation framework with permutation testing **(Figure 4C)**. PLSDA-based visualization also demonstrated clear stratification between ATB and LTBI using these 3 features **(Figure 4D)**.

**Figure 4.**
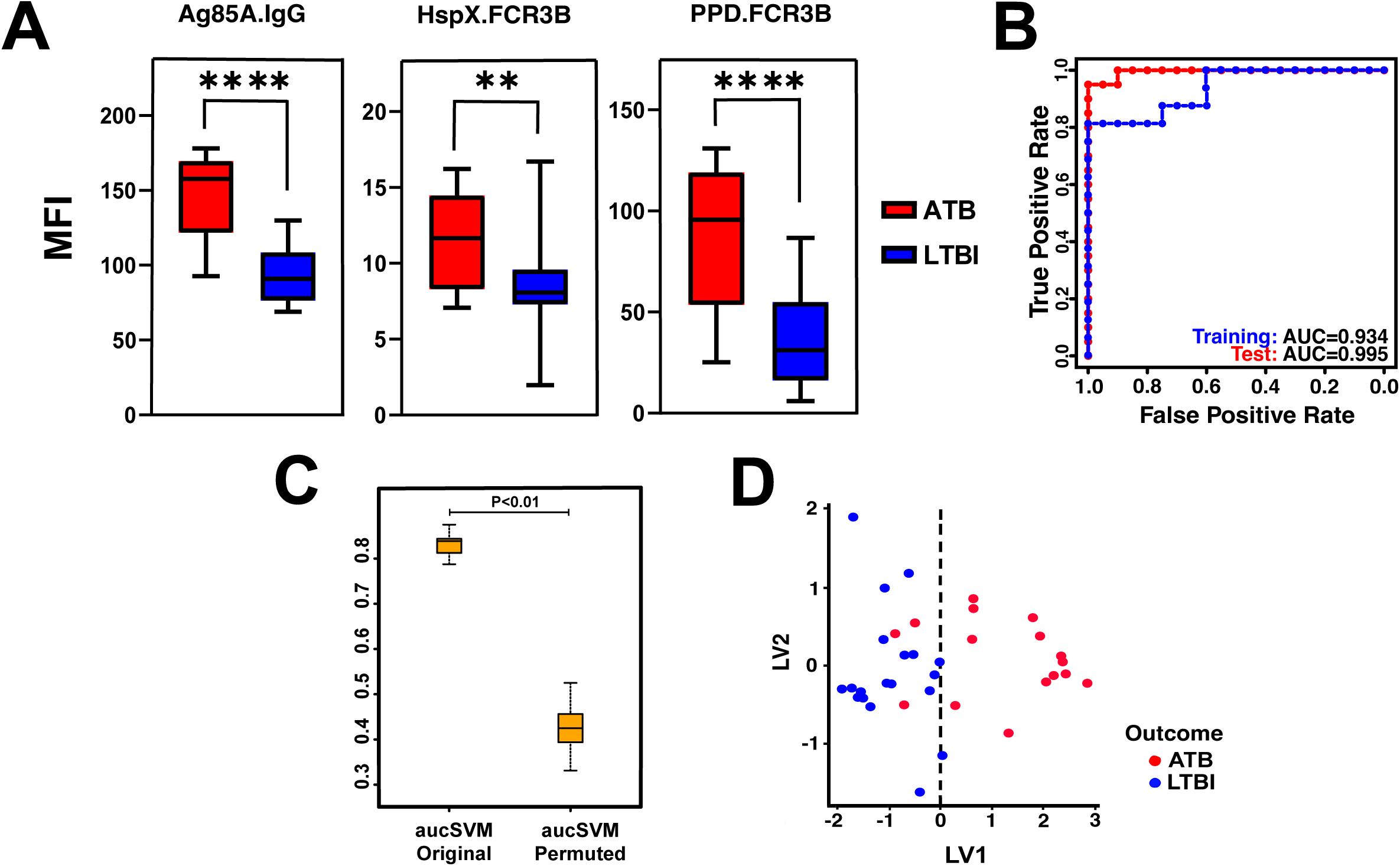
Machine learning-based feature selection using Mtb antibody profiling on a point-of-care diagnostic platform **(A)** Three LASSO-selected features from model built using IgG and FcγR3-binding profiles against 5 Mtb antigen specificities measured on POC platform. **(B)** Receiver Operating Curves (ROC) showing LASSO-SVM model performance using the three features in (A) to discriminate between ATB and LTBI samples in the secondary validation cohort (in red) compared with the performance in the discovery cohort (in blue). **(C)** Control plot showing performance of LASSO model on antibody-omic profiling data consisting of 10 IgG + FcγR3B features from South African cohort sera (n=36) using probe-directed enzymatic metallization POC assay with cellphone-based optical readout. Permuted denotes performance of the model on shuffled data in a matched cross-validation framework (negative control). **(D)** PLS-DA using only the three LASSO-selected features from the model in (A) to discriminate between ATB and LTBI.

To further test the robustness of this biomarker and POC detection approach, beyond the in-cohort cross-validation and permutation testing, we applied the identified biomarker from the South Africa cohort on an independent set of samples from a different cohort as well. This validation cohort (n = 49; **Table. S1, Figure S3C–D)** included individuals with ATB (n=20), LTBI (n=20), and endemic controls (n=9) from South Africa and Vietnam. We tested the performance of the prior trained 3-feature model on this new dataset without any model re-training or data leakage. The model, as trained earlier, showed high diagnostic accuracy (AuRoC>0.9) in this independent testing cohort as well **(Figure 4B, in red)**.

### Multiplexed Sample-Sparing POC Measurement of Mtb-specific Ab Fab and Fc Biomarker

Finally, to further extend ease of use, we developed and demonstrated the ability to measure this multivariate Ab Fab and Fc-based TB biomarker in a multiplexed manner on this POC platform from single sample drops **(Figure 5A).** This was done by harnessing the localized nature of the silver deposition assay, which enables the use of multiple antigen spots placed within a single sample drop. This multiplexed design here was used for 7-plex measurements—covering 5 Mtb antigens plus positive (Protein A/G) and negative (BSA) control antigens—to capture both IgG titer and FcγR3B binding signatures each from single sample drops. The performance of this highly reconfigurable multiplexed biomarker detection platform was demonstrated in the representative image shown in **Figure 5B**, which shows the measurement results from in pooled ATB and LTBI sample from South Africa cohort (n=36). The quantification of the silver darkness in **Figure 5B** is shown in **Figure 5C-D**. Further testing and validation of the multiplexed, sample-sparing format of the assay, using individual samples from this cohort, is shown in **Figure S4** as well.

**Figure 5.**
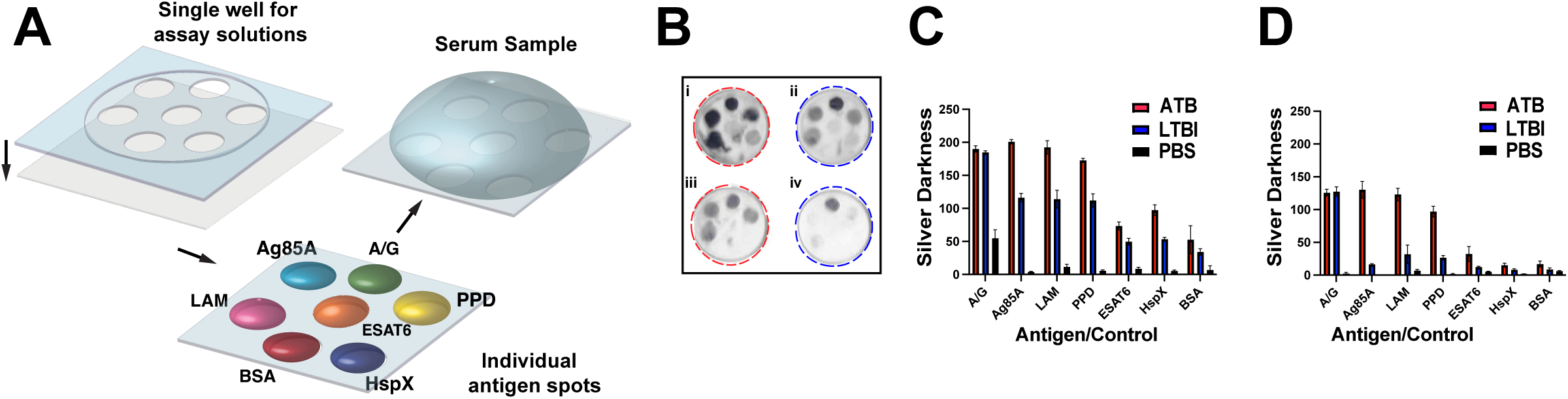
Antigen multiplexed measurement of TB antibody Fc biomarker on a point-of-care diagnostic platform. **(A)** Conceptual overview of antigen multiplexed POC biomarker detection platform. **(B)** Distinct localization of silver deposition of positive control (Protein A/G), negative control (BSA), and 5 Mtb antigens from pooled ATB (in red, n=16) and LTBI (in blue, n=20) plasma from South Africa cohort (n=36) probed with IgG (i-ii) and FcγR3B (iii-iv) respectively. **(C)** Quantification of silver darkness for detection of Mtb-specific IgG antibody response from pooled ATB (in red, n=16) and LTBI (in blue, n=20) plasma from South Africa cohort (n=36) **(D)** Mtb-specific FcγR3B antibody response between ATB, LTBI and positive/negative control from pooled ATB (in red, n=16) and LTBI (in blue, n=20) plasma from South Africa cohort (n=36).

## Discussion

Overall, this work presents the first study to perform comprehensive Ab profiling in TB coupled to machine-learning to define a minimal ATB biomarker, translate it to a multiplexed POC platform and validate it in two independent cohorts (n=85). We found a 5-feature biomarker including antigen-specific Ab titer, FcγR binding and glycosylation that achieved high diagnostic accuracy (AuRoC>0.9). Notably, higher FcγR3B binding to Abs across many Mtb antigens emerged as a leading single Mtb-specific Fc difference in ATB. Based on this, a high avidity multimeric Fc probe based enzymatic silver metallization assay was developed. This was used to adapt the multivariate TB biomarker (titer and Fc) to a low cost yet high sensitivity, multiplexed and portable diagnostic platform. A further optimized 3-feature biomarker (titer and FcγR3B) was established on the POC platform and validated in two independent cohorts. Measuring this biomarker accurately from a single drop of plasma or serum, this work addresses the WHO-defined critical need for non-sputum biomarker-based POC TB diagnostics^2^.

Previous work has characterized Ab Fc profiles in TB and found differences in bulk (non-antigen-specific) Ab glycosylation in ATB vs LTBI. Specifically, higher levels of sialylation and galactosylation were found on bulk IgG in LTBI compared to ATB^14^. Further, differences in two Mtb antigen-specific Abs (PPD and Ag85A) have also been reported with higher sialylation and galactosylation found on these Abs as well in ATB^15^. However, antibody Fc glycosylation measurements have often been limited to characterizing bulk Ab glycosylation or a limited set of antigen-specific Abs due to methodological limitations arising from the high sample requirement of glycoprofiling methods such as MS and CE^26^ compared to relatively low abundance (titer) of Mtb-specific Abs. These methods require the prior isolation of antigen-specific Abs (10-40µg) before enzymatically cleaving off glycans and measuring them. With low abundance Abs, like many of the Mtb antigen-specific Abs, this translates to a high starting sample volume (up to or >1mL serum or plasma per antigen). We have developed a lectin-based glycoprofiling method that enables sample-sparing and highly multiplexed measurements of antibody glycoprofiles directly in a bead binding-based assay^18^ from small sample volumes (<10µL). Using this method, here, we measured the sialylation and galactosylation profiles of 10 Mtb antigen-specific and 2 non-Mtb antigen-specific Abs **(Figure S1).** This revealed differences in sialylation in 4 Mtb-specific Abs and both the non-Mtb-specific Abs measured as well, all of which were found to be higher in LTBI compared to ATB **(Figure 1C).** This corroborates prior Ab glycosylation findings in TB but also provides deeper resolved antigen-specific Ab glycoprofiles than have been available in the context of TB as yet.

In this study, we then built comprehensive antigen-specific Ab-omic profiles including both glycosylation and FcγR binding of Abs. Comparing these Fc measurements revealed that FcγR3B binding to Mtb-specific Abs, but not non-Mtb-specific Abs, was higher in ATB vs LTBI **(Figure 1C).** Further a machine-learning based analysis of these high-dimensional Ab-omic profiles showed that a minimal five feature signature, containing two Mtb-specific Ab FcγR3B binding features (HspX and ESAT6), but not any of the glycan features, was able to accurately discriminate ATB vs LTBI (AuRoC>0.9). This indicates that FcγR3B-binding maybe a more robust antigen-specific biomarker for ATB than Ab titer or glycosylation alone.

FcγR3B is known to be uniquely expressed in humans and is also the highest expressed of any FcγR on any cell type^27^. It is constitutively expressed on neutrophils only which are the most abundant phagocytes in circulation. Neutrophils have been shown to have a dual function in TB contributing to both protection and inflammation-mediated pathology^28^. Interestingly, earlier work, using blood transcriptomics, has also found a neutrophil-driven transcriptional signature for ATB^29^. FcγR3B, while often considered functionally homologous with FcγR3A^30^, has been shown to have both a inhibitory role in antibody-dependent cellular cytotoxicity (ADCC) but also an activating role in other neutrophil functions such as antibody-dependent neutrophil phagocytosis (ADNP) and Neutrophil Extracellular Trap (NET) release. In sequencing studies, higher FcγR3B copy number has been found to be associated with HIV-TB coinfection compared to HIV alone in people living with HIV (PLHIV)^30^. Low copy number of FcγR3B, on the other hand, has been found to be strongly associated increased risk of autoimmune diseases like systemic lupus erythematosus (SLE) and rheumatoid arthritis (RA)^31^. It is also worth noting here that Fc receptor binding of Abs, in general, is known to be modulated by Fc glycosylation of Abs^13^. This is true for FcγR3B as well^32^ but the role of specific glycosylation modifications driving differences in FcγR3B binding, especially in the context of polyclonal immune responses that occur after infection, are as yet unclear. Given the breadth and complexity of these prior findings about FcγR3B, defining the specific functional role of FcγR3B in TB, and its potential modulation by antibody Fc modifications, clearly requires more study. Meanwhile, given its dominance in the multivariate Ab Fc signature found here for ATB, here we next developed methods for the use of FcγR3B-binding of Mtb-specific Abs in a POC diagnostic.

The low natural affinity (K_D_∼10^-5^M) of monomeric FcγR3B^32^ poses a significant challenge for use in simple yet sensitive and robust POC diagnostic assays. In vivo, the clustering of Fcγ receptors on immune cell surfaces is essential for transducing activation signals and driving key effector functions such as phagocytosis, antigen-dependent cellular cytotoxicity (ADCC), cytokine release and complement activation^33^. Multimerization effectively increases the number of receptors binding (avidity), promoting stronger and more stable interactions with Ab-antigen immune complexes. This enhanced avidity not only improves binding strength but also fine-tunes activating and inhibitory signaling pathway interactions, an approach that has also been explored as a potential therapeutic strategy^34^. Inspired by this, we engineered a FcγR3B multimer via the coupling of biotinylated FcγR3B to a polymeric form of streptavidin-labeled HRP. This high avidity multimeric probe provided high sensitivity to detect FcγR3B-binding to Mtb-specific Abs in the POC assay **(Figure 3)** and help distinguish ATB and LTBI samples.

Using a probe-directed enzymatic silver metallization assay on a flexible polymeric substrate then enabled both high-throughput and multiplexed assay formats for inexpensive evaluation of a combined Ab titer and FcγR3B signature from low volumes (<10µL) of samples across two different cohorts. Titer-based TB diagnostics have historically failed in TB, and their use is recommended against by the WHO^35^. Here, we find that this limitation may arise mainly from relying solely on quantity or titer of Abs which can be heterogeneous. We address this fundamental limitation of serological tests by not depending exclusively on titer but instead leveraging the ability of our POC diagnostic platform to measure Ab qualitative Fc features as well. A comprehensive biomarker including Mtb-specific Ab quality, specifically FcγR3B binding, and quantity (IgG levels) was found to achieve high diagnostic accuracy (AuRoC>0.9) across two cohorts here. This is indicative of the value of integrating Ab quantity and quantity as a disease-state discriminative biomarker for TB and beyond^17,18^.

The POC diagnostic platform developed here is based on a simple dry readout of the optical density of enzymatic silver deposition on a flexible polymer substrate. We have shown earlier that this silver readout can be used to optically^22^ or electronically^23–25^ quantitate biomarker concentrations. The localized nature of this silver deposition allows multiplexed measurements from single small volume drops of samples. This assay matches the immunoassay performance of gold-standard ELISAs while offering low sample use, multiplexing **(Figure 5)** as well as high throughput **(Figure 3)** due to the ease of reconfigurability of the polymer substrate. It thus also overcomes the inherent limitations of traditional lateral flow assays (LFAs), which often provide lower sensitivity, semi-quantitative outcomes and lack multiplexability. We note here that despite these critical advantages in the measurement of the multivariate TB biomarker discovered here, the platform used here currently remains more complex to use compared to commercial LFAs as pipetting steps are still involved. This can be overcome in future work via the development of inexpensive automation tools. Also, the biomarker itself can be highly amenable to use in any binding-based assay platform, including emerging multiplexed LFA platforms as well. However, this will require additional future assay development and optimization work.

Additional future work is needed to further assess the robustness of the biomarker reported here in larger cohorts. Further, the evaluation of its performance to detect clinical heterogenous states like subclinical TB will be important as well. Evaluating it in longitudinal cohorts is needed to clarify its utility for therapeutic monitoring and for detecting treatment failure susceptibility (RIF resistance), broadening its impact beyond initial TB diagnosis alone. The inexpensive POC platform developed here, and microfluidic automation tools we have reported earlier, should enable such studies, which can be carried out directly in endemic regions as well, without requiring expensive instrumentation or sample transport to specialized laboratories.

## Conclusion

In summary, we have shown here the development and application of Ab-omics to discover a novel Fc-based biomarker for TB diagnostics and its translation to an inexpensive multiplexed POC detection platform. We observed that Ab titers and several Fc properties differ across ATB and LTBI including most prominently Mtb-specific Ab FcγR3B binding. Thus, a highly sensitive multimeric enzyme-labeled probe to measure FcγR3B binding was developed. A minimal biomarker defined using three Ab features, including Mtb antigen-specific IgG and FcγR3B binding was then measured using microscale enzymatic metallization for POC detection. This showed high diagnostic performance in detecting ATB, across two independent cohorts, achieving AuROCs which meets and indeed surpasses the current WHO targets for POC TB diagnostics and approaches the corresponding standard for lab-based TB diagnostics. This work also establishes antibody-omic profiling as a viable method for disease-state specific Fc-centric biomarker discovery and delivering multiplexed, sample-sparing, inexpensive tests suitable for deployment in resource-limited settings.

## Methods Materials

Tween 20 (97062-332) was obtained from VWR. Deionized water (DIW, LC267405), and phosphate buffer saline (PBS, 21-040-CVR) were obtained from Fisher Scientific. Bovine Serum Albumin (BSA) (A7030) was obtained from Sigma-Aldrich. Tuberculin protein (PPD) was obtained from AJ Vaccines. Purified Ag85A (NR-53525), MTB Lipoarabinomannan (LAM) (NR-14848), CFP-10 (NR-49425), MPT64 (NR-49435), MPT32 (NR-14862), GROES (NR-14861), PSTS1 (NR-53528), ESAT6 (NR-49424) and HspX (NR-49428) Recombinant Reference Standard antigens were obtained from BEI Resources, NIAID, NIH. Poly-HRP conjugated Streptavidin (N200), HRP-conjugated streptavidin (N100) and streptavidin-PE were obtained from Thermo Fisher. Peroxidase AffiniPure Donkey Anti-Human IgG (H+L) (AB_2340495) was purchased from Jackson ImmunoResearch. Biotinylated Human FcgR1, FcgR2A, FcgR2B, FcgR3A and FcgR3B were purchased from Acro Biosystems. PE-labelled mouse anti-human IgG, IgG1, IgG2, IgG3, IgG4, IgA, IgA1, IgA2 and IgM were purchased from SouthernBiotech. EnzMet for General Research Applications (6010-45ML) was obtained from Cedarlane. SNA and RCA lectins were obtained from Vector Laboratories.

### Clinical Samples

Prior bio-banked samples from 85 adult participants from two separate cohorts (South Africa cohort and validation cohort) were obtained (Table S1). The South Africa cohort consisted of plasma samples from 36 individuals from the Cape Town region of South Africa. Further details of the overall cohort have been published earlier^36^. These participants were all >18y of age and seronegative for HIV. The secondary validation cohort consisted of serum samples from 49 individuals with latent TB infection, active TB disease and endemic controls from Vietnam and South Africa, sourced from FIND Diagnostics. These individuals were all >23 y of age and seronegative for HIV. Latent TB infection was defined as patients having a positive Interferon Gamma Release Assay (IGRA; QuantiFERON Gold IT: IFU>=0.35 IU/ml) while active pulmonary TB disease was defined as patients having both/either a positive sputum smear or culture for Mtb. Endemic control participants were defined as having a negative sputum smear or culture for Mtb and negative IGRA.

### Antibody-omic Profiling Assays

The technique used for multiplexed biophysical profiling of antibodies was adapted from methods we have reported earlier^17,18^. Briefly, Luminex MAGPIX magnetic microsphere beads of distinctive regions were coupled to Mtb-specific (Ag85A, LAM, PPD, ESAT6, HspX, MPT64, MPT32, PSTS1 & GROES) and control (Flu & Tetanus) antigens. Coupled beads were then blocked and stored in blocking buffer (1XPBS, 0.1% BSA, 0.1% Tween) at 4° C. These antigen-coupled beads were then pooled and diluted using assay buffer (1XPBS, 0.1% BSA) to form a working bead solution. These beads were then incubated with human plasma (1:1250 dilution) at a sample: bead ratio of 1:9 for 1 hr at 37°C in a flat bottom 96-well plate. Following sample incubation, sample-bound beads were washed three times using wash buffer (PBS, 0.1% Tween) and a magnetic plate separator. Resuspended beads were then exposed for 30 mins to fluorescently labelled probes. PE-labelled mouse anti-human probes at a concentration of 1µg/ml in 1XPBS were used to measure IgG, IgG1, IgG2, IgG3, IgG4, IgA, IgA1, IgA2 and IgM. Antigen-specific FcγR binding profiles were measured using tetramerized FcγR1, FcγR2A, FcγR2B, FcγR3A and FcγR3B at a concentration of 1µg/ml in 1XPBS. FcγR tetramers were formed by reacting biotinylated Fcγ receptors and streptavidin-PE at a 4:1 molar ratio for 20 mins. Antigen-specific glycosylation profiles were measured using PE-labelled lectins. SNA-PE was used to measure sialyation while RCA1-PE was used to measure galactosylation. Following probe incubation, beads were washed three times using wash buffer and resuspended in Luminex drive fluid. The 96-well plate was then read using a Cytek Aurora Flow Cytometer for IgG, IgG1-4, IgA, IgA1-2, IgM, FcγR1, FcγR2A, FcγR2B, FcγR3A and FcγR3B probes and a Luminex MAGPIX instrument for SNA and RCA probes. All assays were performed in duplicate and a correlation coefficient of R2 > 0.8 was verified for technical replicability. An arithmetic mean of the two measured MFI values from the replicates is then used as the readout.

### POC Diagnostic Platform Preparation

Polydimethylsiloxane (PDMS) film (0.1mm thickness, Greene Rubber Company) was laser-cut to create an array of wells with diameters of 2 mm. Another thin piece of PDMS was cut to fit the plane rectangular shape of a typical glass microscope slide (25 mm X 75 mm). The PDMS layers were soaked in 5% Alconox solution then rinsed thoroughly using DI water. Following air drying, scotch tape was used to remove any remaining dust particles. The two layers of clean PDMS were then visually aligned on a clean glass microscope slide with the PDMS layer with the well array being placed onto of the plane PDMS. The two layers of PDMS were secured to the slide using parafilm on either side of the glass slide to prevent leakage.

### Multiplexed Biomarker Detection Assay on POC Diagnostic Platform

Binding assay and readout methods on the POC platform were adapted from methods we have reported earlier^22^. TB antigens (Ag85A, LAM, PPD, ESAT6 and HspX) were prepared at a concentration of 50µg/ml in PBS then added to each well of the prepared PDMS coated slide. Following overnight incubation at 4°C in a humidified chamber, the slide was blocked using blocking buffer (1% BSA in 0.1% Tween 20 in PBS (0.1% PBST)). The antigen coated and blocked slide was then washed using 0.1% PBST, PBS and DI water. After spin-drying the slide, 3µL of diluted plasma or serum sample (1:20 dilution) was added to each well and incubated for 1 hr. Following washing and drying, an IgG probe solution (1:800 dilution of HRP-conjugated IgG) or poly-HRP conjugated FcγR3B (1mg/ml) was added to each well. After another 1hr incubation, washing and drying, equal volumes of silver metallization substrate components A, B and C were sequentially added and incubated for 4, 4 and 5 minutes respectively. The reaction was stopped by quenching the slide in DI water and then drying it. Slides were imaged using either an office scanner (Brother MFC-L2710DW) and readouts obtained using ImageJ. Alternatively, slides are imaged using a cellphone camera and readouts obtained using the MO-BEAM app developed and reported earlier17.

### Statistical Analyses and Machine Learning Models

A two-step machine-learning approach, similar to what has been previously described earlier^17,18^, was used to identify a minimal set of predictive biomarkers from the antibody-omic profile datasets. Specifically, following normalization using z-scoring, feature selection was carried out on the high dimensional dataset (features >> number of patients) using the Least Absolute Shrinkage and Selection Operator (LASSO) L1 regularization method to limit overfitting. Then, classification was carried out on the down-selected features using Support Vector Machines (SVM). Model performance was evaluated using a 10-fold cross validation framework. 500 independent 10-fold cross-validation replicates were performed to account for the different ways in which the training and test folds can be split. Significance of model performance was evaluated using permutation testing where the performance of the selected model was evaluated against a negative control model built with data with randomly shuffled ATBI-LTBI labels, within the cross-validation framework described above. A partial least squares discriminant analysis (PLS-DA) was then carried on the whole dataset using the LASSO-selected features as an independent visualization of the stratification achieved. PLS-DA loading values were also obtained to further explore the relative contribution of the features to the stratification. All the above analyses were performed in R. For univariate analyses, statistical significance was assessed using a Mann-Whitney U test. Correction for multiple comparisons was done using a Benjamini-Hochberg correction, using GraphPad Prism.

## Data Availability

All datasets generated are available upon reasonable request from the corresponding author.

## Code Availability

All code used for analysis and instructions for use to reproduce the results are available are available upon reasonable request from the corresponding author.

## Ethics and Inclusion

This study uses bio-banked samples derived from a cohort established as part of a prior completed and published study where further details of the overall cohort have been described earlier^36^. The study was conducted in accordance with the principles expressed in the Declaration of Helsinki. All participants gave written informed consent for the study, which was approved by the institutional review boards at the University of Cape Town (Protocol# 288/2008) and Emory University (Protocol# IRB00061334). Safe handling of these clinical samples for assays was governed by the biosafety protocols established, approved by and available from the Biological Materials Safeguards Committee (BMSC Permit: B-970-B-001) at the Georgia Institute of Technology. Local investigators (co-authors here: C.D., L.S. & M. d. K) from the So1u1th African Tuberculosis Vaccine Initiative (SATVI) were included throughout the research process. Roles and responsibilities of all investigators were defined through joint planning meetings, ensuring equitable collaboration and capacity-building and training for local researchers. Data ownership, authorship, intellectual property and benefit sharing measures were established through mutual institutional agreements. Relevant local and regional TB research is cited^36^.

## Data Availability

All data produced in the present study are available upon reasonable request to the authors

---

**Figure S1.**
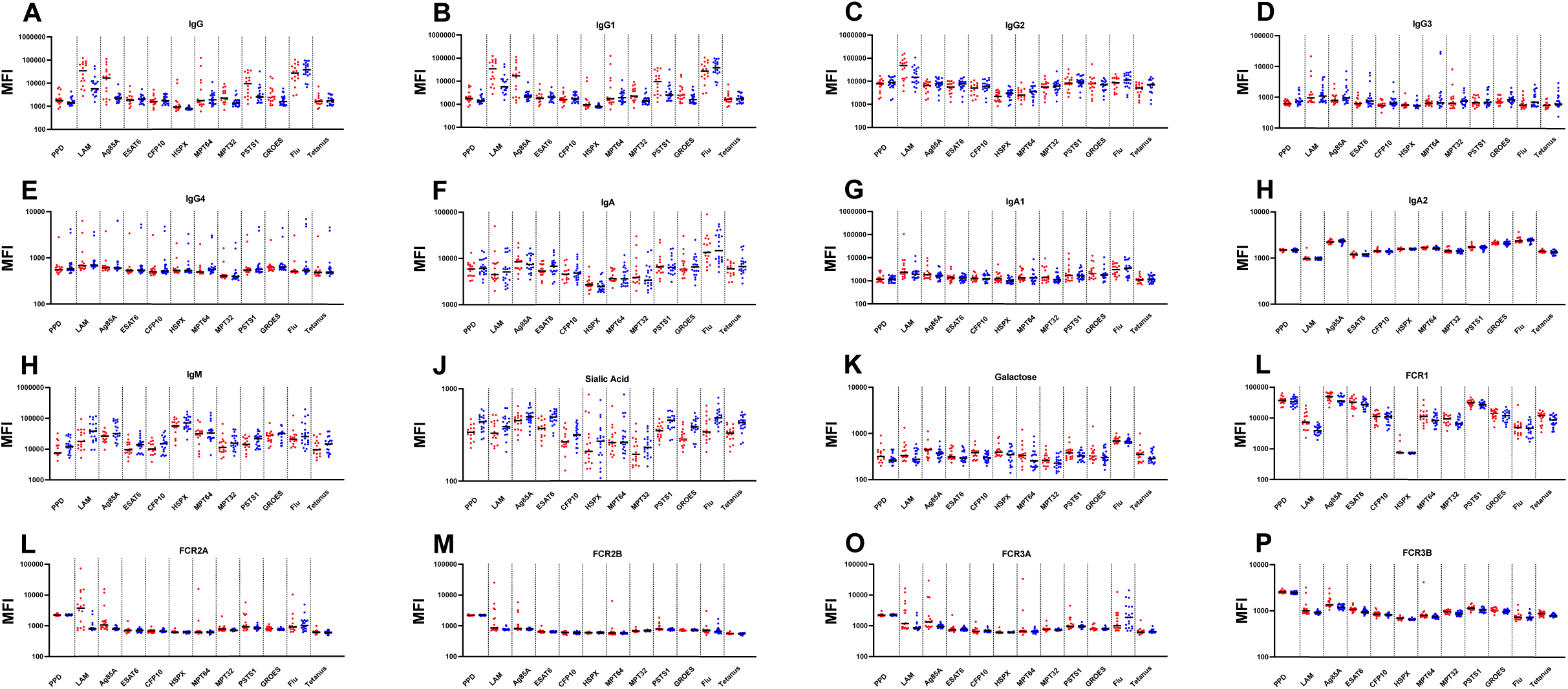
(A-P) Plots of Mtb-specific and non-Mtb antigen-specific antibody features measured in ATB (in red, n=16) and LTBI (in blue, n=20) plasma from the South Africa cohort (n=36) using multiplexed bead-based antibody-omics platform.

**Figure S2.**
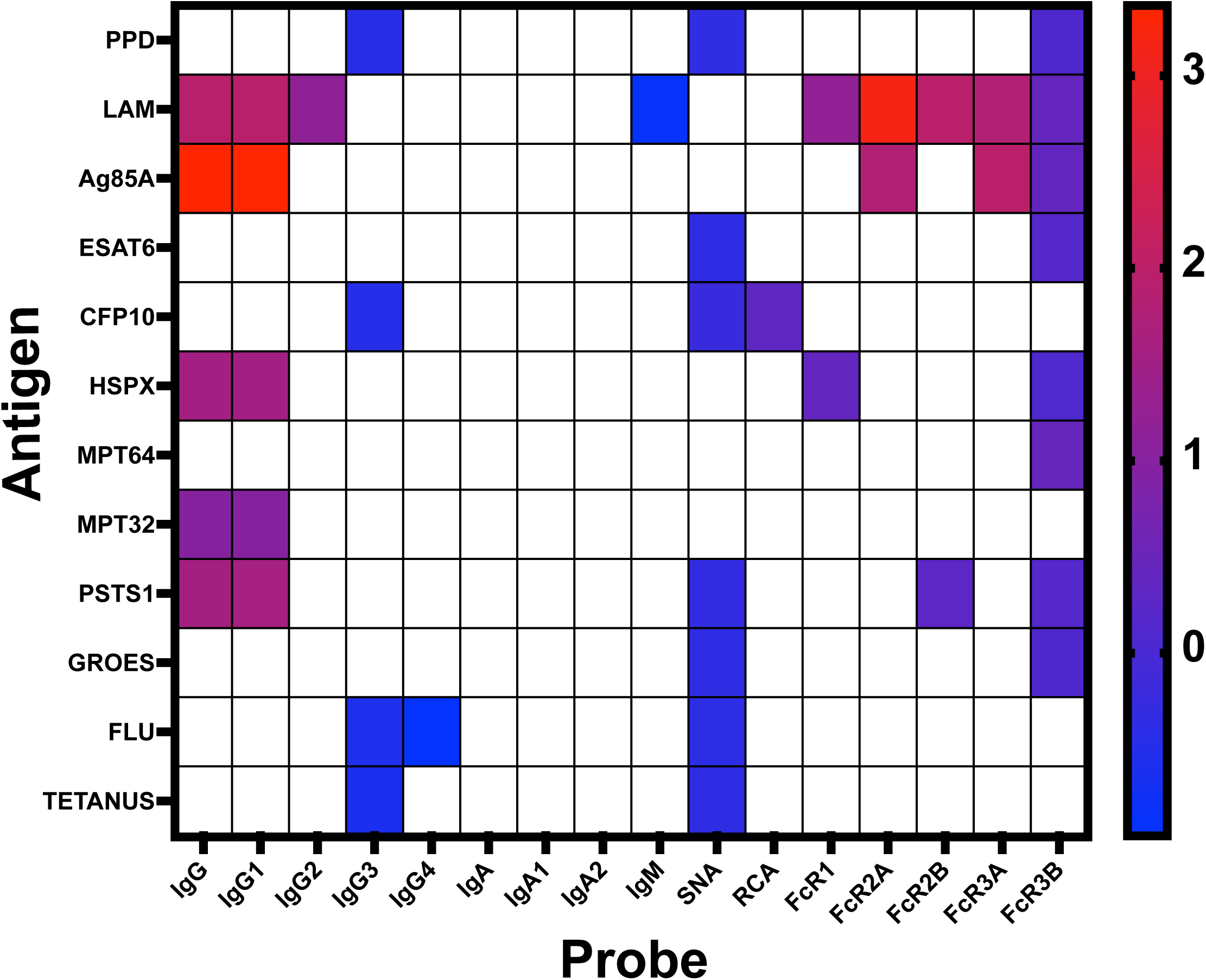
Univariate analysis of Mtb-specific and non-Mtb antigen-specific response in ATB and LTBI plasma from the South Africa cohort (n=36). **(A)** Heatmap showing Log2 fold change of antibody features. Different antigen specificities (Fab) are shown in different rows while different Fc probes are shown in different columns. Only those features which show statistically significant differences (p<0.05) are shown in color (blue increased in LTBI; red increased in ATB). A colormap scale bar for the log2-fold-change is shown on the right.

**Figure S3.**
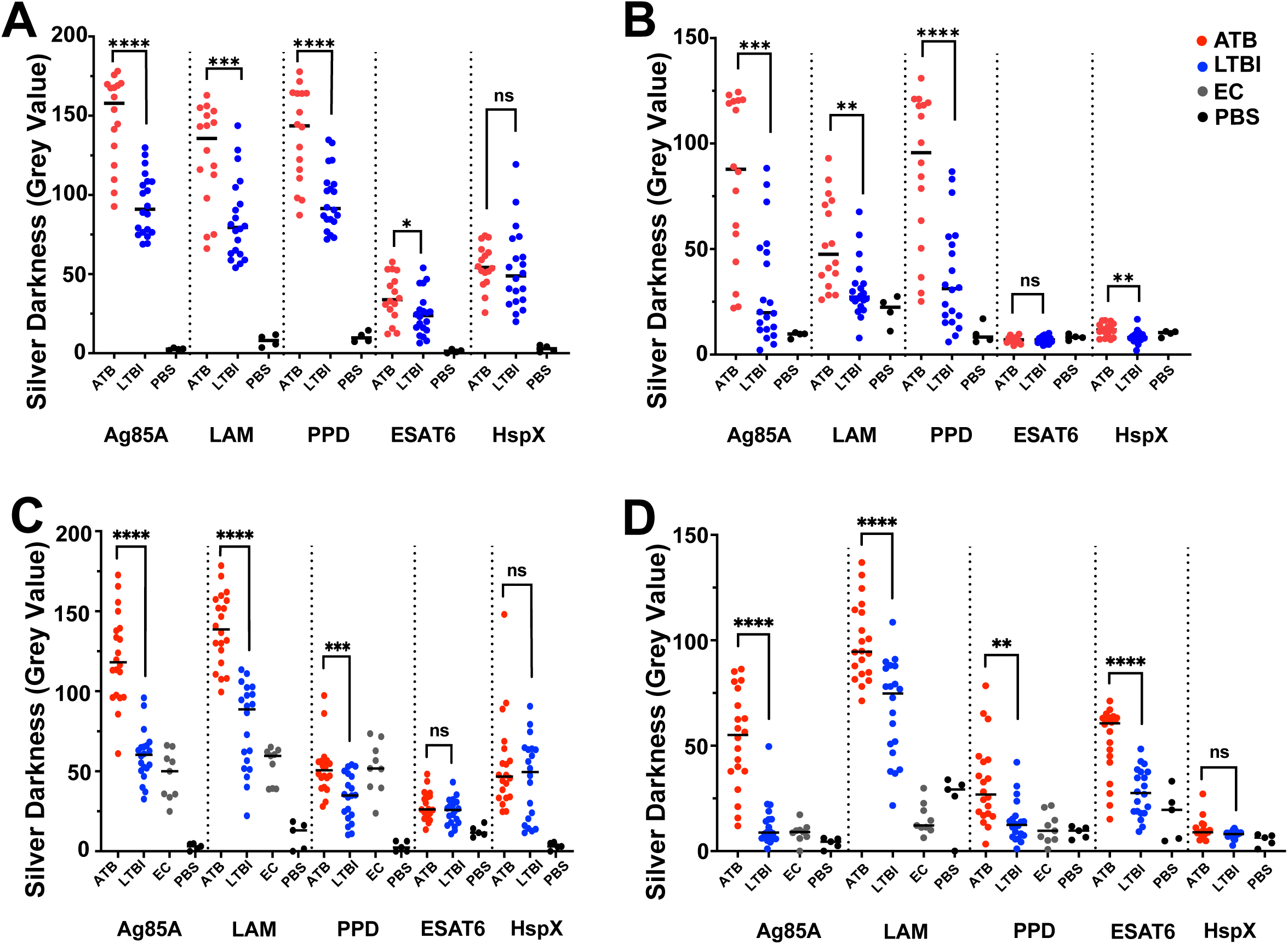
Univariate plots from South Africa and secondary cohorts graphing the Mtb-antigen-specific IgG and FcγR3B response of individual ATB (in red), LTBI (in blue), endemic control (EC; in grey) samples and PBS control (in black) using the point-of-care diagnostic platform **(A)** Quantification of silver metallization response of ATB (n=16), LTBI (n=20) and PBS control shown in with IgG probe in the South Africa cohort **(B)** Quantification of silver metallization response of ATB (n=16), LTBI (n=20) and PBS control shown in with multimeric FcγR3B probe in the South Africa cohort **(C)** Quantification of silver metallization response of ATB (n=20), LTBI (n=20), EC (n=9) and PBS control shown in with IgG probe in secondary validation cohort **(D)** Quantification of silver metallization response of (n=20), LTBI (n=20), EC (n=9) and PBS control shown in with multimeric FcγR3B probe in secondary validation cohort

**Figure S4.**
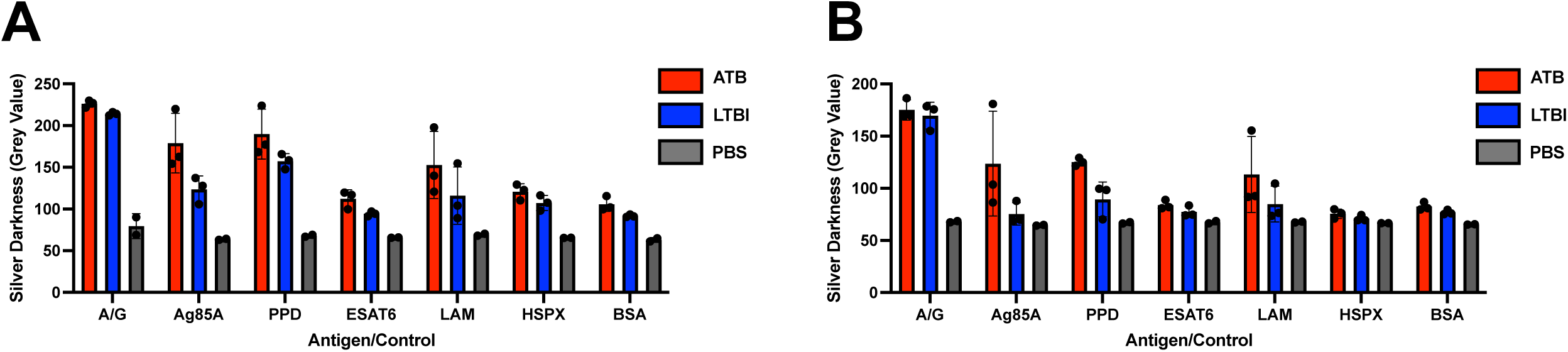
Antigen multiplexed measurement of TB antibody Fc biomarker on a point-of-care diagnostic platform on individual patients from South Africa cohort (A) Quantification of silver darkness for detection of Mtb-specific IgG antibody response from ATB (in red, n=3) and LTBI (in blue, n=3) plasma samples (B) Quantification of silver darkness for detection of Mtb-specific FcγR3B antibody response from ATB (in red, n=3) and LTBI (in blue, n=3) plasma samples.

**Table S1.** Cohort details of training and validation cohorts.

| Cohort Name | Variable | ATB<br>N=16 | LTBI<br>N=20 |  |
| --- | --- | --- | --- | --- |
| South Africa Cohort | Age,years,median (IQR) | 30 (18,51) | 27 (19,50) |  |
|  | Female, n(%) | 8 (50%) | 11 (55%) |  |
|  | Male, n(%) | 8 (50%) | 9 (45%) |  |
|  | HIV positive, n(%) | 0 (0%) | 0 (0%) |  |
|  | Diagnostic Tests |  |  |  |
|  | Culture positive, n(%) | 16 (100%) | - |  |
|  | Smear positive, n(%) | 0 (0%) | - |  |
|  | Interferon Release Assay (IGRA) positive, n(%) | - | 20 (100%) |  |
| Cohort Name | Variable | ATB<br>N=20 | LTBI<br>N=20 | EC<br>N=9 |
| Validation Cohort | Age,years,median (IQR) | 50 (27,80) | 42 (23,71) | 43 (35, 57) |
|  | Female, n(%) | 11 (55%) | 8 (40%) | 8 (89%) |
|  | Male, n(%) | 9 (45%) | 12 (60%) | 1 (11%) |
|  | HIV positive, n(%) | 0 (0%) | 0 (0%) | 0 (0%) |
|  | Diagnostic Tests |  |  |  |
|  | Culture positive, n(%) | 20 (100%) | 0 (0%) | 0 (0%) |
|  | Smear positive, n(%) | 20 (100%) | 0 (0%) | 0 (0%) |
|  | Interferon Release Assay (IGRA) positive, n(%) | - | 20 (100%) | 0 (0%) |

